# Estimating the Burden of Influenza on Daily Activity at Population Scale Using Commercial Wearable Sensors

**DOI:** 10.1101/2021.05.06.21255083

**Authors:** Aziz Mezlini, Allison Shapiro, Eric J. Daza, Eamon Caddigan, Ernesto Ramirez, Tim Althoff, Luca Foschini

## Abstract

The severity of viral infections can vary widely, from asymptomatic cases to complications leading to hospitalizations and death. Milder cases, despite being more prevalent, often go undocumented and their public health impact unaccounted for. We estimated the burden of influenza-like-illness (ILI) by leveraging the widespread use of commercial activity trackers. Analysing data from 15,382 US participants who reported ILI symptoms during the 2018–2019 flu season (before the COVID-19 pandemic) and who had high-density wearable sensor data at symptom onset, we estimated an overall nationwide reduction in mobility equivalent to 257 billion steps lost due to ILI symptoms. This finding reflects significant changes in routines, mobility, and employment and is equivalent to 15% of the active US population becoming completely immobilized for 1 day. Moreover, ∼60% of this impact occurred among individuals who sought no medical care, who would otherwise be invisible to healthcare and public health reporting systems. We validated our measure against self-reported measures of disease severity. We believe this method has applications for public health, healthcare, and clinical research, from estimating costs of lost productivity at population scale, to measuring effectiveness of anti-ILI treatments, to monitoring recovery after acute viral syndromes, such as during ‘long COVID’.

---

More than a year into the COVID-19 pandemic, SARS-CoV-2 has infected more than 131 million people worldwide and caused close to 3 million documented deaths^1^. However, the global burden of the pandemic is suspected to be much higher, as milder infections, the most prevalent, are more likely to go unreported. This type of underreporting is not specific to COVID-19; it applies to other viral infections such as influenza-like illness (ILI).

ILI is highly prevalent in the U.S. and the rest of the world. Every year, ILI is among the leading causes of mortality, severely affecting the youngest and the eldest^2^. Even beyond the severe cases, however, ILI places a considerable burden on society. Previous work has measured this burden in terms of quality-adjusted life years, financial cost, work productivity, and/or absenteeism^3,4^, but these measures often rely on self-reported estimations of varying quality. Furthermore, an important portion of that burden might remain unobserved because milder ILI cases can be underrepresented or overlooked in studies that recruit participants solely from clinical care institutions or that analyse medical records or claims data^5^.

Commercial wearable sensors provide unique opportunities to understand objective behavioural and physiological changes associated with respiratory infections such as ILI and COVID-19 in real-world settings. Participants who consent to data sharing can allow the unobtrusive and continuous collection of rich data without the burden of manual, subjective, self-reported input regarding the severity of disease. By their nature of being continuously produced over time (such as hour-level or day-level step counts), these data can complement measures of total disease burden to study the dynamics of the burden over the course of the disease^6^. Indeed, previous studies have shown the ability of wearable data to track ILI-associated behavioural and physiological changes in activity (such as resting heart rate, sleep, and step counts)^7,8^, reflect state-wide ILI prevalence^9^, and even forecast future ILI events at the individual level based solely on abnormal activity data^10^.

We used large-scale wearable data to quantify the burden of ILI at a national level. Specifically, we used a cohort corresponding to a large population from whom both wearable data and frequent ILI survey data were collected. We first present a method for assessing ILI burden at the individual level and then reweight the resulting estimates to reflect the US population using estimates of flu burden published by the US Centers for Disease Control and Prevention (CDC)^11^. Our aim was to assess whether the burden of ILI increases with markers of increased disease severity (medical visits, hospitalization, symptoms) and explore associations between covariates and the ILI burden trajectory from onset to recovery.

## Results

### Participants

After applying the selection criteria, demographic information, daily steps data from wearable sensors, and ILI survey data were available for a total of 15,382 participants. On average, participants were 31 years old; 77% were women, 11% were men, and 12% were nonbinary or chose not to report their gender; 87% were white; and 92% had a minimum of a high school diploma (Table 1). For their ILI event, 19% of participants sought medical attention, and only 55 (0.4%) were hospitalized.

**Table 1.** Demographic and clinical characteristics of the participants.

|  |  |  |  |  |
| --- | --- | --- | --- | --- |
| Characteristic | *n* | Overall (*N* = 15,382) | Female* (*n* = 11,850) | Male (*n* = 1727) |
| Median age (IQR) | 13,871 | 31 (26–38) | 31 (26–38) | 33 (28– 39) |
| Age category, *n* (%) | 13,577 |  |  |  |
| 18–29 |  | 5061 (37) | 4562 (38) | 499 (29) |
| 30–39 |  | 5433 (40) | 4652 (39) | 781 (45) |
| 40–49 |  | 2085 (15) | 1778 (15) | 307 (18) |
| 50–59 |  | 829 (6.1) | 714 (6.0) | 115 (6.7) |
| 60+ |  | 111 (0.8) | 98 (0.8) | 13 (0.8) |
| 65+ |  | 58 (0.4) | 46 (0.4) | 12 (0.7) |
| Race/Ethnicity, *n* (%) | 13,525 |  |  |  |
| Asian or Pacific Islander |  | 554 (4.1) | 430 (3.6) | 124 (7.2) |
| Black or African American |  | 490 (3.6) | 425 (3.6) | 65 (3.8) |
| Hispanic or Latino |  | 666 (4.9) | 550 (4.7) | 116 (6.7) |
| White |  | 11,815 (87) | 10,399 (88) | 1416 (82) |
| Education, *n* (%) | 13,456 |  |  |  |
| Did not finish high school |  | 86 (0.6) | 77 (0.7) | 9 (0.5) |
| High school graduate/GED |  | 981 (7.3) | 872 (7.4) | 109 (6.4) |
| Some college, no degree |  | 2,802 (21) | 2,467 (21) | 335 (20) |
| Trade/technical/vocational school |  | 570 (4.2) | 500 (4.3) | 70 (4.1) |
| Associate or bachelor's degree |  | 6310 (47) | 5496 (47) | 814 (48) |
| Graduate degree |  | 2408 (18) | 2082 (18) | 326 (19) |
| Doctoral degree |  | 299 (2.2) | 250 (2.1) | 49 (2.9) |
| Medical, *n* (%) | 13,536 |  |  |  |
| Attended |  | 2603 (19) | 2,332 (20) | 271 (16) |
| Unattended |  | 10,933 (81) | 9,483 (80) | 1,450 (84) |
| Hospitalization, *n* (%) | 13,563 |  |  |  |
| Hospitalized |  | 55 (0.4) | 48 (0.4) | 7 (0.4) |
| Not hospitalized |  | 13,508 (100) | 11,790 (100) | 1718 (100) |
| *Sex information was missing for 1805 participants. The median age (IQR) for this group was 32 (26–38). |  |  |  |  |

### Baseline modelling

We trained the linear mixed model on all data except the ILI periods and the matching control periods. We then used the model to predict the step count for each individual for each day of the ILI and control periods.

Fig. 1 shows the mean differences between the predicted and observed step counts. As expected, the step differential averages were near-zero for control periods and the days before symptoms in the ILI period (indicating typical healthy behaviour). In contrast, the step differential averages deviated substantially from zero during the symptomatic ILI periods, showing the expected abnormal behaviour during the ILI event. The differences in step differentials between case and control periods were statistically significant only for Days 0–10 of symptom onset, with the step differentials that corresponded to an ILI event being negative and lower than those corresponding to control periods. The average effect of an ILI event was thus a reduction in the number of steps on Days 0–10 of symptom onset. There was no significant difference between the groups during Days −4 to −1 (the latent phase) or on Day 11 and beyond.

**Fig. 1.**
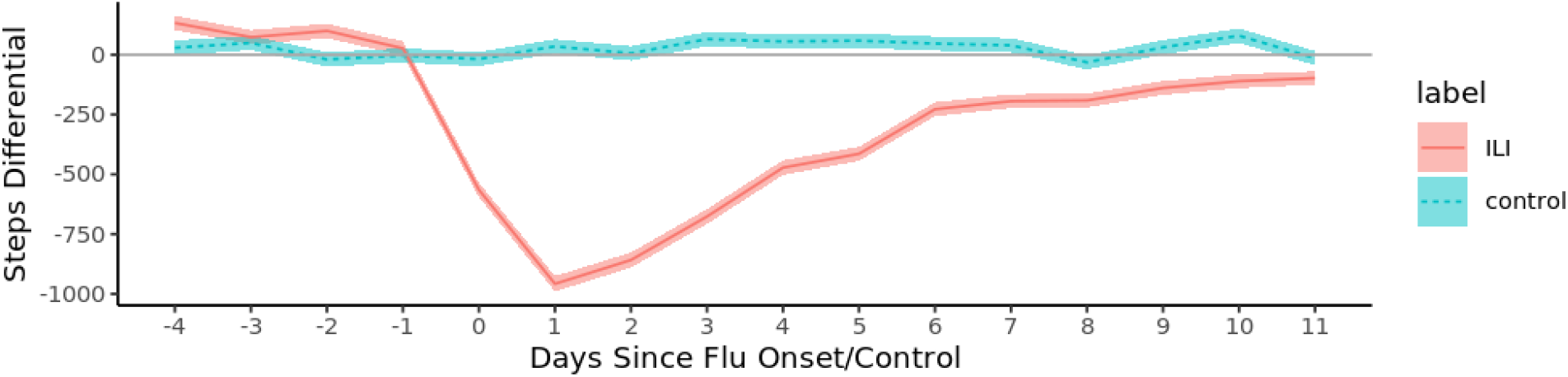
Average step differentials. The mean and standard error during ILI events (red) and matched control periods (in blue). The step differentials are the average across participants of the per-participant difference between the predicted counterfactual step counts and the observed step counts.

Despite the strongly discernible effect on steps overall, there was a great deal of heterogeneity (Fig. 2): 47% of participants showed little to no decrease (<1000 steps lost) or an overall increase in steps during Days 0–10 of their ILI event. This proportion was 59% for the control periods. However, when we focused on only the examples with >15,000 steps lost, such cases represented 20.8% of the ILI examples and only 10.6% of the control examples.

**Fig. 2.**
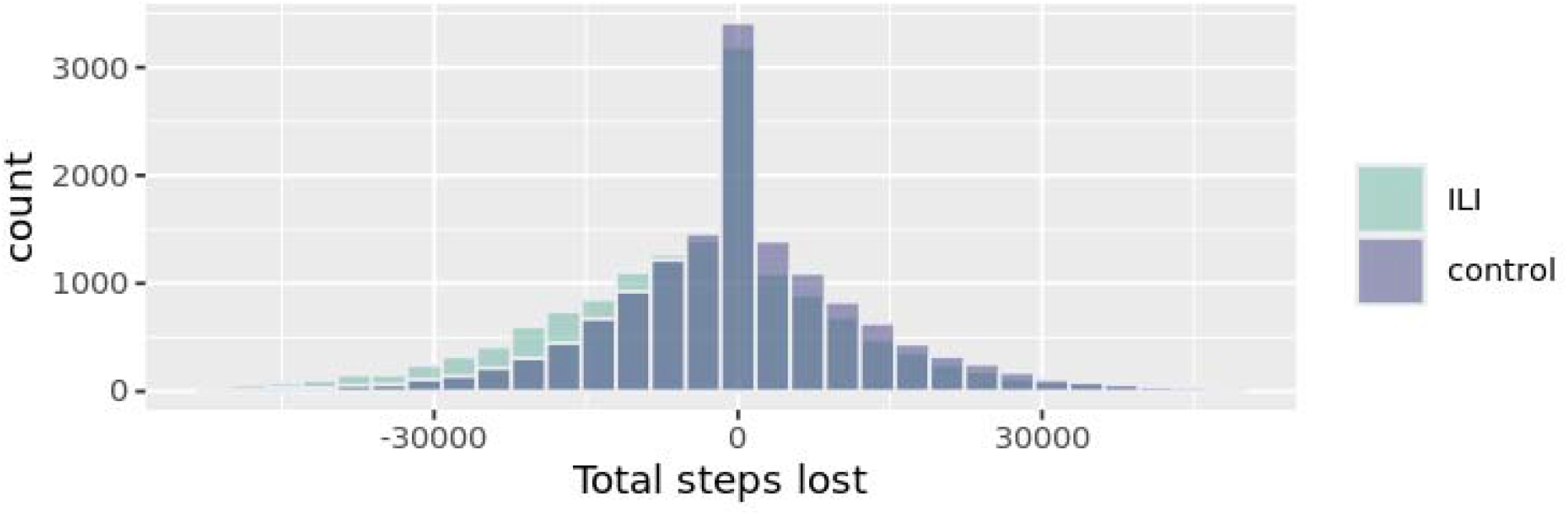
Histogram of total ILI burden across participants measured in equivalent of lost steps. Total steps lost during Days 0–10 during ILI events (green) and matched control periods (in blue).

### Overall ILI burden

Across our sample, an average 956 steps (95% confidence interval [CI], 892–1020) were lost on the worst day of the ILI event, corresponding to Day 1 after symptom onset (see Fig. 1). In comparison, the loss estimation during the control periods did not change significantly (35 steps lost, 95% CI, −23–93). Over the course of the ILI (Days 0–10), the average cumulative loss was 4487 steps (95% CI, 4192–4782). For control periods, the burden was much smaller, with 347 steps lost (95% CI, −613 – −80, *P*=0.092 after false discovery rate [FDR] multiple hypothesis correction).

We reweighted the sample using self-reported age, sex, race/Hispanic ethnicity, and educational attainment to approximate the overall US population 18 years and older. Using this approach, we estimated that a total of 257.6 billion (95% CI, 225.2–290.1 billion) steps were lost during ILI events in the U.S. during the study period (Table 2). The corresponding analysis for the matched control periods yielded no significant change (13.4 billion steps lost, 95% CI, −9.4–36.2 billion).

**Table 2.** Estimated total burden and maximal burden on Day 1 of ILI, in terms of steps lost, with 95% confidence intervals (in billions of steps, B) for the whole US population.

|  |  |  |
| --- | --- | --- |
|  | Total Days 0–10 | Day 1 Maximal burden |
| Unattended | -153.2B (-177.6B – -128.9B) | -27.2B (-31.9B – -22.5B) |
| Attended, not hospitalized | -100.5B (-110.8B – -90.3B) | -13.5B (-15.6B – -11.4B) |
| Hospitalized | -3.9B (-4.5B – -3.2B) | -0.6B (-0.9B – -0.3B) |
| Overall ILI events | -257.6B (-290.1B – -225.2B) | -41.2B (-47B – -35.5B) |
| Overall control periods | -13.4B (-36.2B – 9.4B) | -2B (-10.4B – 6.4B) |

Of the 257 billion steps lost due to ILI, 59.4% (153.2 billion steps, 95% CI, 177.6–128.9) reflected individuals who had not sought medical attention.

### Association of covariates with ILI burden

In both univariate tests and logistic regression analyses, neither sex nor race/ethnicity had an association with ILI burden, whereas age and education level did. As shown in Fig. 3, the ILI step burden increased with age and with the level of education. The latter might be explained by reduced job precarity among the more highly educated and the ability to take days off to recover from home leading to a larger reduction in the number of steps taken.

**Fig. 3.**
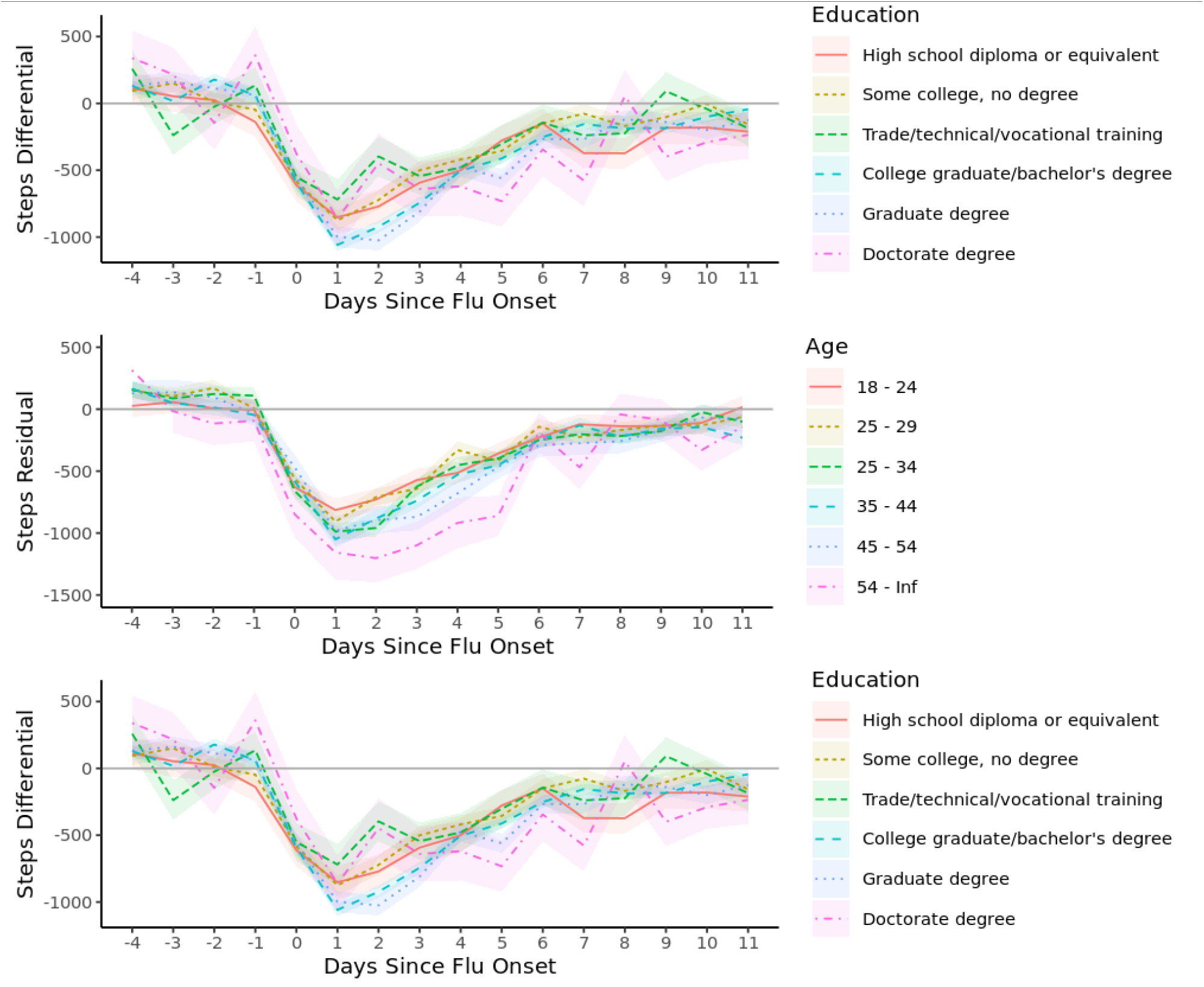
Average step differentials between participants with ILI and controls by education level and age group. Mean and standard error are shown for each group.

Participants who were seen by a physician had a higher flu burden compared with the rest of the sample (Fig. 4). This would be expected, as more severe ILI episodes likely led to requests for medical assessment. This was true whether the flu diagnosis was positive or negative. Similarly, participants who reported being hospitalized had a higher burden, although the standard error was large given the small number of reported hospitalizations (*n =* 55).

**Fig. 4.**
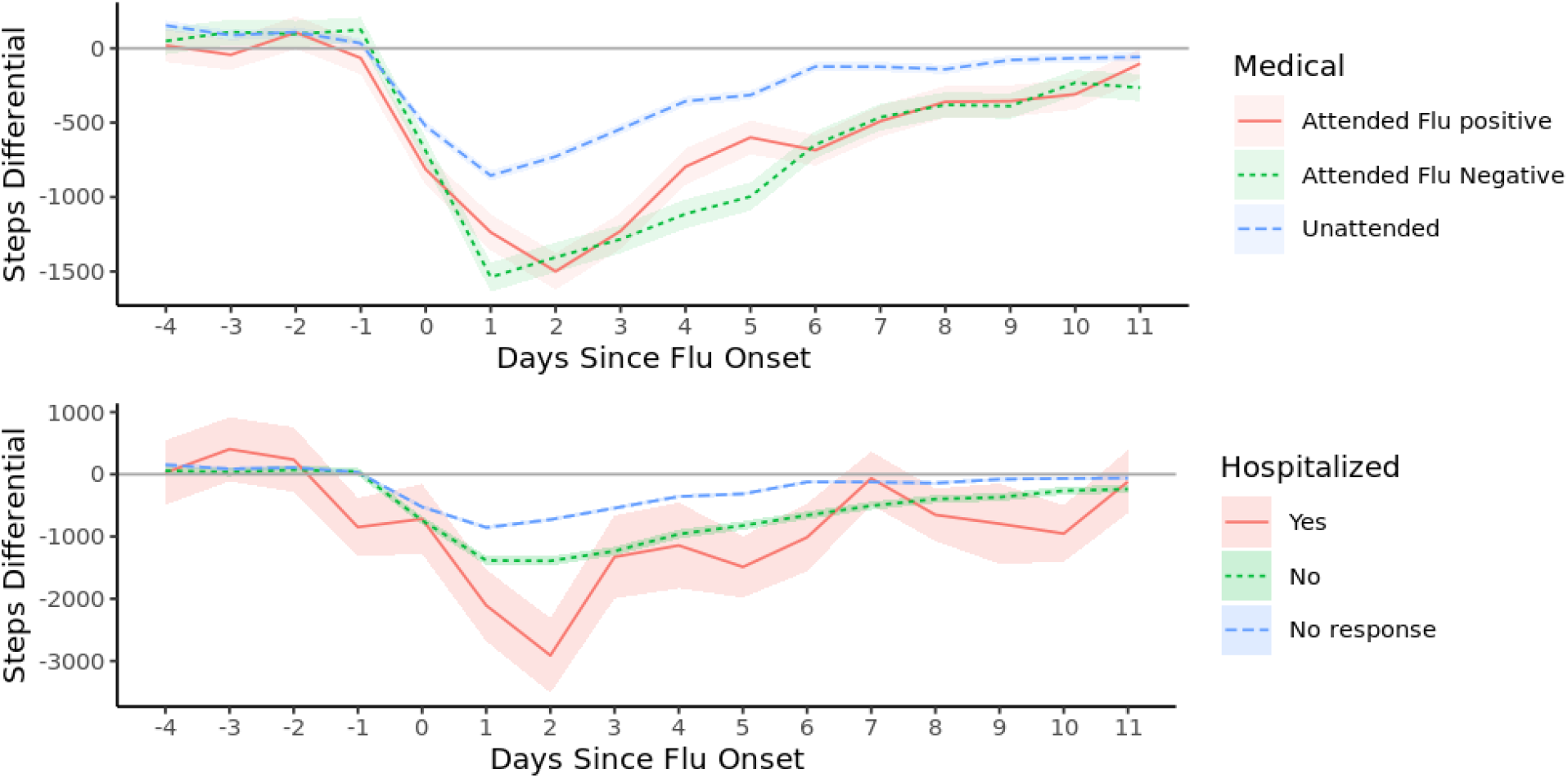
Average step differentials between participants with ILI and controls by medical care received (unattended, attended, and hospitalized). Mean and standard error are shown for each group.

ILI burden was also associated with both the number of workdays missed and self-reported overall health (Fig. 5). As expected, participants who took more time off lost more steps than those who reported working through their illness. This can be interpreted in two ways: either participants with milder disease presentation did not need to take time off work, or using steps to measure the ILI burden underestimates illness severity among participants who are less inclined to take time off because of socioeconomic reasons. Participants who self-reported ‘very good’ to ‘excellent’ health also lost more steps. This is likely because their higher baseline activity before ILI led to a larger difference between the baseline and ILI states.

**Fig. 5.**
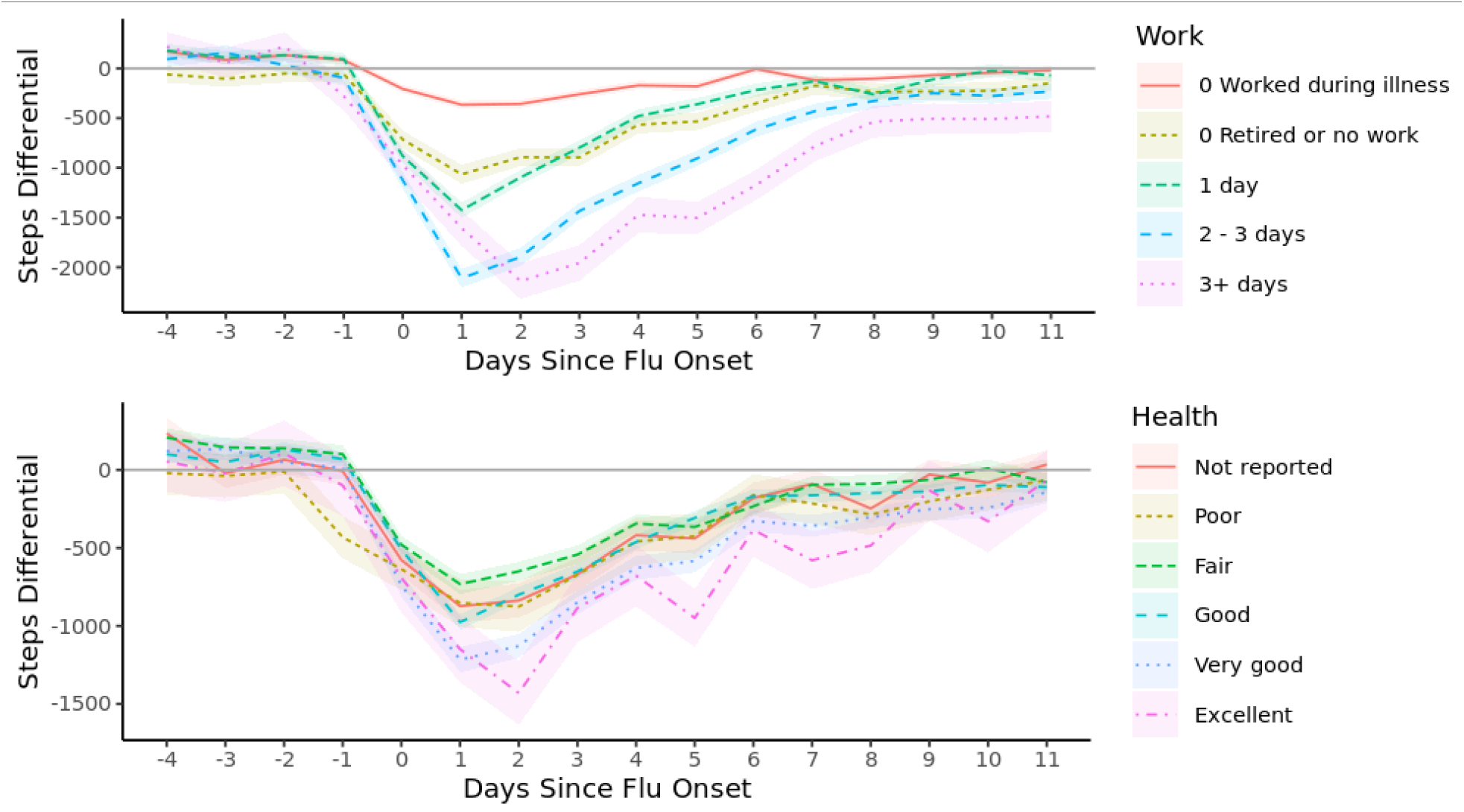
Average step differentials between participants with ILI and controls by general health status and number of days off work. Mean and standard error are shown for each group.

Finally, we investigated the association of different symptoms with the trajectory of ILI burden. We grouped symptoms into three sets of similar associations. Fig. 6 shows the associations of fever, chills, and body aches with steps lost during ILI events (Days 0–10). Each of these symptoms corresponded to greater step loss throughout the event.

**Fig. 6.**
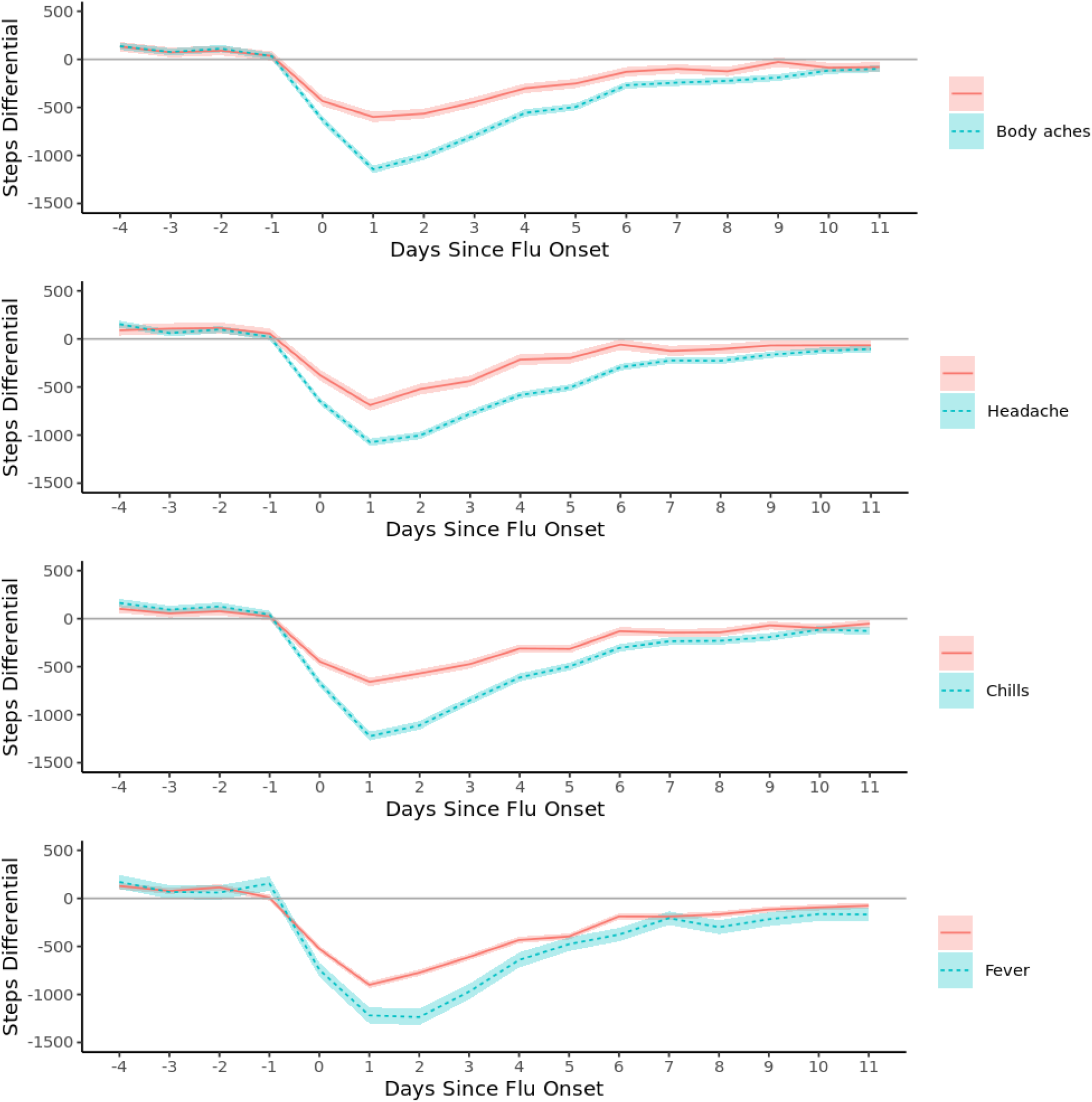
Average step differentials between participants with ILI and controls by symptom experienced. Mean and standard error are shown for each group.

In contrast, nausea and diarrhoea symptoms were associated with a higher burden on Days 0–2 but not afterward (Fig. 7).

**Fig. 7.**
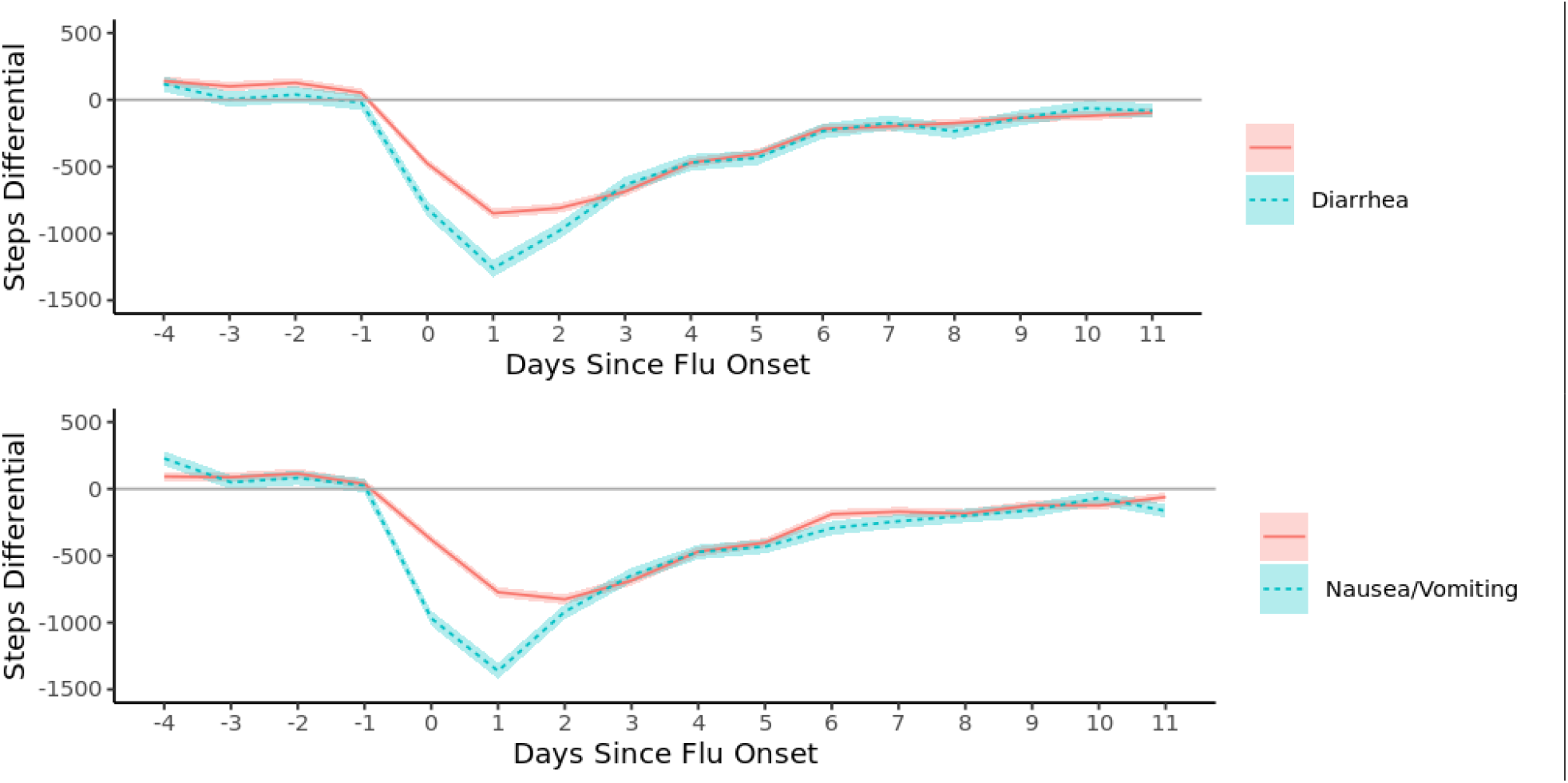
Average step differentials between participants with ILI and controls by symptom experienced. Mean and standard error are shown for each group.

The final set consisted of respiratory symptoms (coughing, shortness of breath, and sore throat), each of which yielded different associations on burden trajectory (Fig. 8). Participants with sore throat had a lower burden (fewer steps lost) in the initial phase of the ILI (Days 0–2) but converged to the same level of steps differential in the remaining days compared with participants who did not report sore throat. Similarly, participants with cough exhibited a lower burden on average on the first 2 days; however, they exhibited a higher burden on Days 3–10 and a slower recovery compared with participants without a cough. Finally, participants who reported shortness of breath showed a similar average burden on Days 0–2 but a higher burden on Days 3–10.

**Fig. 8.**
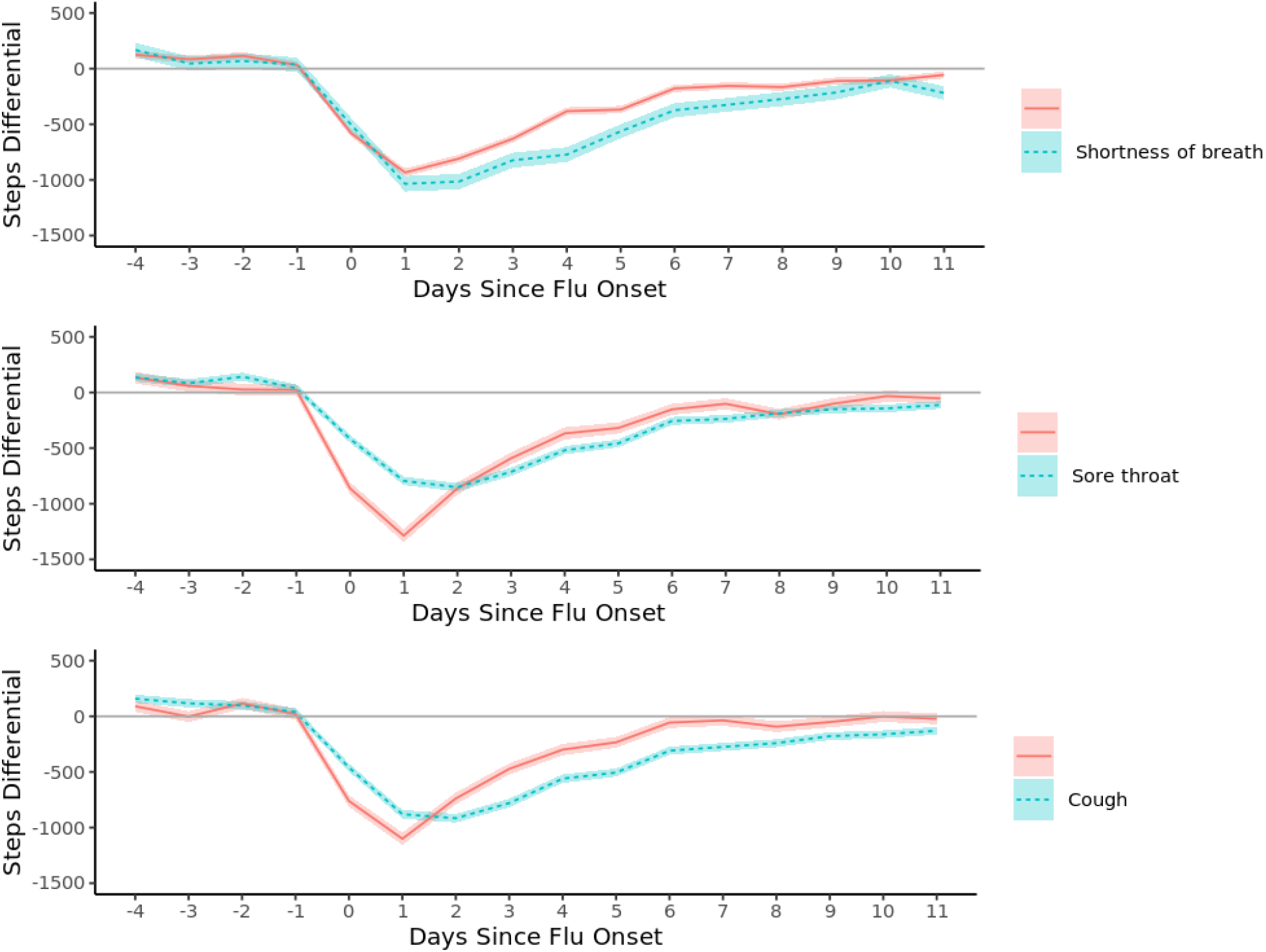
Average step differentials between participants with ILI and controls by symptom experienced. Mean and standard error are shown for each group.

## Discussion

We present an approach for quantifying the population burden of ILI, based on changes in daily activities obtained from wearable sensors linked to survey responses from the 2018–2019 flu season. Our results show that wearable sensors can be used to estimate the burden of ILI at the individual and national levels in terms of the number of steps taken daily. We additionally investigated how different demographic covariates might modify this burden, and how different symptoms associate with different trajectories of burden throughout the course of the disease.

Our study is not without limitations. The measure of burden used in this paper was the average number of steps lost. This is a directly quantifiable, objective measure with available data collected by wearable sensors. However, this measure of burden could be considered an imperfect proxy for more interesting but less quantifiable measures, such as loss of quality of life and loss of productivity due to ILI. For example, a proportion of our participants did not lose a meaningful number of steps and did not take time off work despite the ILI. For such participants, it is difficult to assess the real burden of ILI in terms of productivity or quality of life. The steps differential is not likely representative of the full disease burden in this scenario: Not losing steps does not mean that there was no burden on the participants.

A potential improvement on this work in the future would be to refine our baseline model predicting step counts. We plan to explore the use of deep learning and of individual-level models taking into account auto-correlation to potentially achieve less noisy predictions of baseline steps. A better estimate of activity in the absence of ILI will allow us to better quantify the effects of ILI.

A similar approach to that taken in this paper could be used in a variety of use cases. In public health settings, this measure of burden developed could be used to measure the impact of other viral infections, such as COVID-19, and to monitor over time the effectiveness of policies and interventions to reduce their spread. In healthcare settings, the same approach could also be applied to quantifying the burden of other chronic diseases, including COVID-19 post-viral syndrome (‘long COVID’) or seasonal disorders. Finally, with adequate controlling for confounding factors, the approach could be used in clinical research to measure effectiveness of therapies.

## Methods

This study received expedited review and independent institutional review board (IRB) approval from Solutions IRB (Yarnell AZ, protocol ID #2019/07/18). This IRB waived the requirement for informed consent from participants due to the retrospective design of the study.

### Data collection

The Achievement application (Evidation Health, Inc., San Mateo, CA) is a mobile consumer platform that encourages users to develop healthy habits—such as walking, meditating, and logging meals—and provides incentives for them to participate in research by completing surveys and sharing their data from commercial-grade wearable sensors^12–14^. Since 2017, the application has been used for voluntary monitoring of annual waves of influenza virus infections^15^.

We developed an ILI survey and administered it during the 2018–2019 flu season. (See Supplemental Information for the survey instrument.) Every 2 weeks, surveys queried members as to whether they had had any flu-like symptoms in the preceding 14 days. Members who reported symptoms were then asked to describe specific symptoms they experienced, symptom onset date, recovery date (if available), healthcare-seeking behaviour, diagnosis of influenza, lost workdays, household illness, and vaccination history days.

We also collected minute-level step data from October 2018 through June 2019 from participants who agreed to linking their Fitbit sensor data to the Achievement platform. Minute-level activity time series were transformed into day-level time series per user, indicating the total number of steps daily. Several quality-control criteria were applied to ensure that analyses included only participants using Fitbit who had dense activity data coverage throughout the flu season (particularly the days surrounding the ILI) and only participant-days with sufficient sensor wear time (see the data density criteria in the Supplemental Information).

The survey also captured the demographic characteristics of age, sex, race/Hispanic ethnicity, and education level achieved.

### Statistical analysis

Our dataset enabled robust characterization of the time course of behaviour during ILIs (Fig. 9). Participants’ self-reported descriptions of ILI events (survey responses) were paired with their activity time series, passively recorded from their wearable fitness tracker. ILI onset dates were used to index the activity time series, with flu Day 0 marking self-reported symptom onset. There were three time periods of interest:

**Fig. 9.**
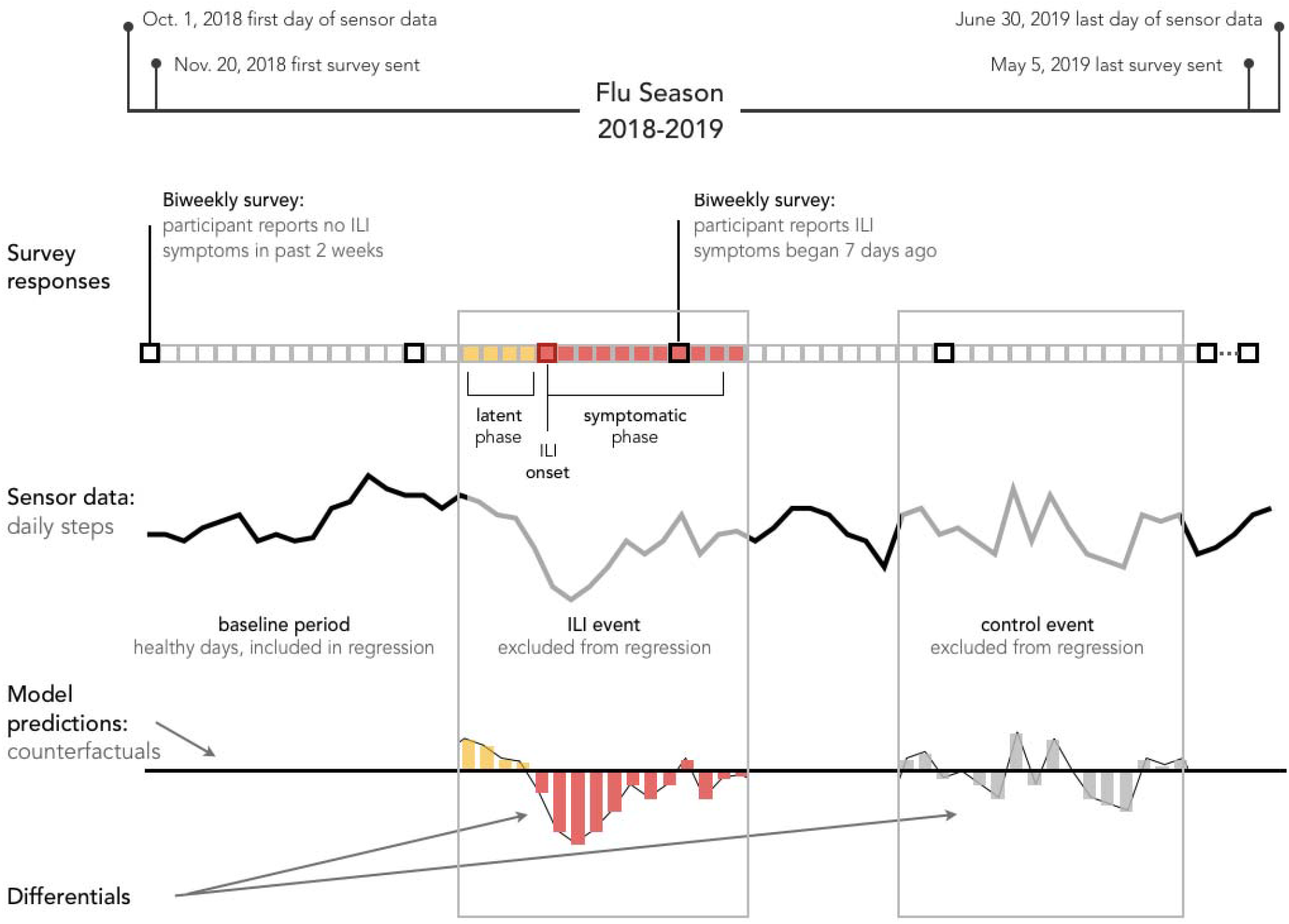
Overview of data collection during the study period, with a sample graphical representation of change in steps during an ILI event.

1. The latent phase: ILI Days −4 through −1. The incubation period of the flu is 1–4 days, according to the CDC^16^.
2. The symptomatic phase: ILI Days 0–11. This choice was based on our data, where the last day with a statistically significant^17,18^ change in steps was noted on Day 10.
3. The baseline or healthy phase: the remaining days in the time series.

Additionally, we created matching control events. Each individual had one flu event and one control event whose dates corresponded to the flu event of another randomly sampled participant. This ensured that the distribution of control event dates matched the distribution of ILI dates.

We fitted a longitudinal multilevel regression model (LME4 package for R^19^) to days during the baseline period. The model was trained on all data excluding flu and control periods (Days −4 to 11 around each event). This baseline model aimed to quantify typical healthy behaviour by accounting for both group-level trends (e.g., on average, people walk less during winter months than in the spring and fall) and individual differences between participants (e.g., weekly step activity patterns differ between office workers and hospital nurses). Fixed effects were specified for each week of the year separately (as a categorical variable), which captured seasonal behavioural patterns at the group level. We also specified random effects (participant level) for a 5-degree polynomial fitting the week of the year and a random intercept (participant level) for each day of the week to account for individual differences in weekly activity patterns.

Next, the baseline model was used to predict each participant’s behaviour for *all* days of the flu season, including all three time periods of interest. Note that multilevel models make unique predictions for each individual. The predictions obtained are analogous to counterfactuals^20^, i.e., the number of steps for each individual if they had not had an ILI event. We then computed step differentials as the difference between the observed activity and the predicted activity for each participant-day. These step differentials corresponded to the effect of the ILI for each participant-day. We were interested in the average step differentials across participants. These are akin to the average treatment effect (ATE) in the causal inference literature^21^. The baseline model mean absolute error (MAE) was 2375 steps per day for the baseline period it was trained on, and 2477 for the unseen control periods. The statistical significance of differences in step differentials between cases and controls was assessed by means of the Wilcoxon signed rank test, the *t*-test, and a novel statistical test for heterogeneous effects^22^.

We also calculated the step differentials between the ILI and control periods (Days −4 to 11) per individual. These allowed us to estimate the ILI burden on each individual in our data for each day and to verify that the baseline model predictions contained no observable bias. We could also assess any potential association of covariates such as age, sex, race/Hispanic ethnicity, and disease symptoms with ILI burden.

Because our sample of participants was not representative of the overall US population—i.e., it was not a random probability sample, and differed notably from the US population in distributions for sex and age, for example—we attempted to adjust our quantification of flu burden through poststratification. We reweighted our sample to match the relative proportions and absolute size of the US population by age group, sex, race/Hispanic ethnicity, and educational attainment, as reported by the US Census^23^.

For each day of the flu period, we used a linear mixed model with the step differentials as outcomes and the demographic factors used for restratification (age, sex, race/Hispanic ethnicity, educational attainment), along with the CDC’s disease severity levels (attended medical care, hospitalized) as predictors. Using this model, we could predict the ILI burden for each group or stratum corresponding to every possible combination of covariates. Within each group, the burden was summed over flu Days 0–10 (the only days showing statistically significant differences between cases and controls). These burden predictions were then reweighted by the US Census estimates of population size per demographic category and CDC estimates of ILI prevalence among demographic groups, and finally summed to create an estimate of the total US ILI burden. This procedure of estimating the national ILI burden was repeated on the control periods for comparison, and 95% confidence intervals were estimated by bootstrapping (100 times). (See the Validation section of the Supplemental Information for additional validation details.)

Finally, we tested whether any demographic covariates—such as age category, sex, race/ethnicity category, education level category, medical attention (Y/N), and hospitalization (Y/N)—were associated with the daily or total ILI burden. We fitted a regression model with the daily or total burden as an outcome and these covariates as predictors. We also performed univariate tests and visually inspected the association of each variable separately.

## Data Availability

The datasets analysed in this study are not publicly available but can be shared for scientific collaboration by contacting the corresponding author and subject to meeting requirements of the data use and protection policy of the institution.

## Acknowledgments

This work was supported by Evidation Health, Inc. The authors thank Catherine Wang, Benjamin Bradshaw, Andrew Trister, Kevin Konty, and Yuedong Wang for their scientific guidance, and manuscript and analysis reviews.

