## Supplemental File for "Estimating the Burden of Influenza on Daily Activity at Population Scale Using Commercial Wearable Sensors"

**Supplemental Information**

*Data Density Criteria*

The case definition for influenza-like illness (ILI) is not standard across health organizations^1,2^. We relied on participants’ self-reporting of ILI with no further selection based on symptoms such as fever, as in our previous work^3^. The US Centers for Disease Control and Prevention (CDC) concedes that because of their selection criteria (fever and sore throat or cough), their estimates are a lower bound on the real number of ILI cases and that milder cases may not be captured^4^. Our estimates likely included a broader spectrum of ILI symptom presentations than the CDC’s, given that we retained ILI cases with no fever. From the CDC page^5^: ‘It’s important to note that not everyone with flu will have a fever’.

We chose to consider only one ILI event per participant, corresponding to the first ILI in the season. Some of our participants reported more than one event, with some reported ILIs occurring close together, which could represent repercussions of the initial illness. We inferred ILI events by merging multiple surveys from the same participant with date ranges

encompassing symptom onset and recovery that overlapped or were separated by no more

than 2 days. Then we selected only the first ILI event per participant.

Our choice to ignore multiple events per participant was for simplification purposes, especially when matching with CDC data. This simplification means that our current burden estimates are likely a lower bound on the actual burden of ILI.

To select participants with sufficient data, we followed **data density** criteria. First, valid **days** were defined as those with 10 or more hours of sensor wear time^1^. Then the **data density** criteria were:

1. Baseline period: at least 10% of each weekday needed to be valid.
2. Latent period: at least 1 day during Days -4 through -1 needed to be valid.
3. Onset: Day 0 needed to be valid.
4. Early symptomatic: at least 1 day during Days 1–3 must have been valid (5) overall. Symptomatic: > 50% (6 days) of all days from Days 0 through 9 must have been valid.
5. Participants missing steps data for more than half of the days in the study were excluded.

*Validation*

To validate how representative our estimates of the US population were after poststratification, we estimated ILI prevalence among our reweighted sample and compared it with the prevalence of symptomatic flu-like illnesses across the U.S. in the 2018–2019 flu season. This is estimated annually by the CDC along with estimates of flu-like symptom prevalence, flu-related medical visits, hospitalizations, and deaths. Table 1 compares our estimates based on the Achievement flu survey and the CDC estimates for different age groups.

**Table 1**. Estimates (95% confidence intervals) of the burden of ILI according to the CDC and as derived from the Achievement flu survey, by age category

| **Symptomatic Illness** | | |
| --- | --- | --- |
| **Age** | **CDC Estimate** | **Achievement Estimate** |
| 18–49 | 11,913,203 (10,077,523–16,032,899) | 33,134,570 (32,412,751–33,790,482) |
| 50–64 | 9,238,038 (6,582,690–15,759,286) | 12,066,141 (10,889,546–12,760,730) |
| 65+ | 3,073,227 (2,008,898–6,030,701) | 4,137,900 (3,004,397–4,786,853) |
| **Medical Visits** | | |
|  | **CDC Estimate** | **Achievement Estimate** |
| 18–49 | 4,407,885 (3,498,694–6,064,550) | 6,644,789 (6,335,336–6,963,073) |
| 50–64 | 3,972,356 (2,712,868–6,886,487) | 3,435,232 (2,606,978–3,939,138) |
| 65+ | 1,721,007 (1,097,482–3,394,980) | 1,111,699 (515,191–1,659,903) |
| **Hospitalizations** | | |
|  | **CDC Estimate** | **Achievement Estimate** |
| 18–49 | 66,869 (56,565–89,993) | 229,261 (189,183–275,487) |
| 50–64 | 97,967 (69,808–167,123) | 17,228 (2067–41,402) |
| 65+ | 279,384 (182,627–548,246) | 23,632 (0–62,987) |

Overall, our estimates did not differ greatly from the CDC estimates for medical visits. However, we had a larger number of reported symptomatic illnesses than estimated by the CDC. This is especially true for the younger portion of the population (ages 18–49), where the estimate from our data was much larger. This can be explained by a difference in the definition of ILI used (stricter CDC definition based on symptoms such as fever versus self-reported ILI by Achievement participants). Our sample likely had more cases of milder disease reported.

Of note, we had only 55 total hospitalizations in our study, which explains the large uncertainty in the estimates for that group. Our estimate for hospitalizations in the 18–49 age group is much larger than the CDC estimate. A possible explanation for this discrepancy is mentioned on the CDC page^1^: “The rates provided are likely to be an underestimate as influenza-related hospitalizations can be missed if testing is not performed.”

### Achievement Flu Survey 2018-19 Flu Season

### General Setup

The general setup will be similar to the 2018 flu survey: users will be asked via a one click offer once every two weeks whether they had experienced ILI within the previous two week period. If they respond no, they will be asked again two weeks later. If they respond yes, they will be given the option to complete a survey asking about the specifics of the incident.

#### Point Values

- One click: 5 points / 1-click (sent once every two weeks)
- Survey: 50 points / survey (completed if user reports ILI)

#### One click potential wording

Header: 1-Click Question about the Flu

1st one-click body: We are looking to understand how behavior is related to the flu. Have you recently experienced flu-like symptoms?

- Yes
- No

All following one-click body: We recently asked you if you had experienced flu-like symptoms and wanted to see if anything has changed. Have you experienced flu-like symptoms in the past two weeks?

- Yes
- No

Feed description: Answered 1-click about the flu

Hide Stats?: Yes

#### Survey offer potential wording

Header: Tell us more about your flu-like symptoms

Body: We are looking to better understand how behavior is related to the flu. Answer a brief survey about your flu-like symptoms and earn 50 points!

Call to Action: Take Survey

Feed description: Completed Survey about Flu-Like Symptoms

### Disclosure

### This brief survey will ask you about your recent reported flu-like illness. We’re interested in learning more about the relationship between flu-like illness and activity. You will earn 50 points for completing this survey. Not only will you be able to earn rewards, but you’ll help us set the stage for future research that could benefit millions of people.

### Additionally, if you choose to participate, we may look at your survey responses in conjunction with up to one year of your most recent activity data. We will never share your name or individual responses with anyone. Your responses may also be used to help match you with future research or related opportunities where you can contribute and earn rewards!

### ILI Survey

1. What day did you *first begin to experience symptoms?* If you don’t exactly recall, try your best to select the date that corresponds to when you began feeling ill. (calendar date selection)
2. What was the first date you felt you had completely recovered from your illness? If you don’t exactly recall, try your best to select the date that corresponds to when your symptoms had completely gone away. If you have not yet fully recovered from your illness, please select today’s date. (calendar date selection)
3. As of today, do you feel that you have completely recovered from your illness?
   a. No
   b. Yes
4. When you were sick, did you have a high fever (temp > 102)?
   a. Yes (if yes => proceed to 5)
   b. No (if no => proceed to 6)
   c. I don’t know/Can’t remember
5. How long did your fever persist?
   a. I’m still experiencing fever
   b. < 24 hours
   c. > 4 days
   d. 1 - 2 days
   e. 3 - 4 days
   f. I don’t know/can’t remember
6. What other symptoms did you have? Please select all that apply
   a. Diarrhea
   b. Body aches
   c. Sore throat
   d. Cough
   e. Chills
   f. Nausea/Vomiting
   g. Shortness of breath
   h. Headache
   i. Other (please specify)
7. Did you see a physician or seek medical attention for this illness?
   a. Yes (if yes => proceed to 8)
   b. No (if no => proceed to 12)
8. Did the physician give you a diagnosis of influenza?
   a. Yes (if yes => proceed to 9)
   b. No (if no => proceed to 10)
   c. I don’t know / I can’t remember
9. What was your diagnosis based on?
   a. Nasal swab/sputum test
   b. Blood test
   c. No test
   d. I don’t know/can’t remember
   e. Other (please specify)
10. Were you hospitalized as a consequence of your illness?
    a. No
    b. Yes
    c. I don’t know / I can’t remember
11. Were you prescribed Tamiflu (oseltamivir) or Xofluza (baloxavir marboxil) to treat or manage your symptoms?
    a. I was prescribed Xofluza (baloxavir marboxil)
    b. I was prescribed Tamiflu (oseltamivir)
    c. I was not prescribed either of these medications
    d. I don’t know / can’t remember
12. Have any members of your household (other than yourself) experienced flu-like illness this flu season?
    a. Yes (if yes => proceed to 13)
    b. No (if no => proceed to 14)
    c. I live alone
13. How many members of your household, by age group listed below, have experienced flu-like symptoms during this flu season? If no household member in your household experienced symptoms within an age group please enter 0. [numeric entry]
    a. Number of household members 0-4 years old experiencing flu-like symptoms
    b. Number of household members 5-17 years old experiencing flu-like symptoms
    c. Number of household members 18-49 years old experiencing flu-like symptoms
    d. Number of household members 50-64 years old experiencing flu-like symptoms
    e. Number of household members 65+ years old experiencing flu-like symptoms
14. Did you miss work due to your illness?
    a. No, I worked during my illness
    b. I missed one 1 day of work
    c. I missed 2-3 days of work
    d. I missed more than 3 days of work
    e. Retired and/or work days don’t apply to me
    f. I don’t know / I don’t remember
15. Did you receive the flu vaccine (sometimes called the flu shot) this flu season?
    a. No
    b. Yes
    c. I don’t know / can’t remember
16. Did you receive the flu vaccine last flu season? (September 2017 - March 2018)
    a. Yes
    b. No
    c. I don’t remember
17. Please select the statement below that describes whether you typically get a flu shot (or another form of flu vaccine).
    a. I never have gotten a flu shot
    b. I rarely get a flu shot
    c. I get a flu shot every year
    d. I sometimes get a flu shot
